# Validation of acute myocardial infarction (AMI) in electronic medical records: the SPEED-EXTRACT Study

**DOI:** 10.1101/2020.12.08.20245720

**Authors:** Aldo Saavedra, Richard W. Morris, Charmaine S. Tam, Madhura Killedar, Seshika Ratwatte, Ronald Huynh, Christopher Yu, David Z Yuan, Michelle Cretikos, Janice Gullick, Stephen T. Vernon, Gemma A. Figtree, Jonathan Morris, David Brieger

## Abstract

**Objectives:** To determine whether data captured in electronic medical records (eMR) is sufficient to serve as a clinical data source to make a reliable determination of ST elevation myocardial infarction (STEMI) and non-ST elevation myocardial infarction (NSTEMI) and to use these eMR derived diagnoses to validate ICD-10 codes for STEMI and NSTEMI.

**Design:** Retrospective validation by blind chart review of a purposive sample of patients with a troponin test result, ECG record, and medical note available in the eMR.

**Setting:** Two local health districts containing two tertiary hospitals and six referral hospitals in New South Wales, Australia.

**Participants:** *N* = 897 adult patients who had a hs-troponin test result indicating suspected AMI.

**Primary outcome measures:** Inter-rater reliability of clinical diagnosis (κ) for ST-elevated myocardial infarction (STEMI) and Non-ST elevated myocardial infarction (NSTEMI); and sensitivity, specificity, and positive predictive value (PPV) of ICD-10 codes for STEMI and NSTEMI.

**Results:** The diagnostic agreement between clinical experts was high for STEMI (κ = 0.786) but lower for NSTEMI (κ = 0.548). ICD-10 STEMI codes had moderate sensitivity (Se = 88±6.7), very high specificity (Sp = 99±0.7) and high positive predictive value (PPV = 91±6). NSTEMI ICD-10 codes were lower in each case (Se = 69±6.4, Sp = 96.0±1.5, PPV = 84±6).

**Conclusions:** The eMR held sufficient clinical data to reliably diagnose STEMI, producing high inter-rater agreement among our expert reviewers as well as allowing reasonably precise estimates of the accuracy of administrative ICD-10 codes. However the clinical detail held in the eMR was less sufficient to diagnose NSTEMI, indicated by a lower inter-rater agreement. Efforts should be directed towards operationalising the clinical definition of NSTEMI and improving clinical record keeping to enable an accurate description of the clinical phenotype in the eMR, and thus improve reliability of the diagnosis of NSTEMI using these data sources.

**Article Summary:** *Strengths and limitations of this study:*

- Expert chart review provided a robust evaluation of the reliability and sufficiency of data directly extracted from the EMR for the diagnosis of AMI
- Computational interrogation and extraction of the eMR (via SPEED-EXTRACT) allowed us to use a wide selection for inclusion in the sample on the basis of clinical data *independent of ICD-10 code*, enabling the capture of missed cases (i.e., uncoded AMI) and so determine estimates for the false negative rate and sensitivity
- Results were necessarily based on the subset of patients with sufficient clinical data in the eMR. Inferences from this subset to the wider patient pool will be biased when the availability of records varies with diagnosis
- At least two sources of uncertainty in the gold reference standard we used are indistinguishable: uncertainty due to poor clinical detail in the eMR, and uncertainty due to a weak operational definition of the diagnosis (e.g., NSTEMI).

## Introduction

Acute myocardial infarction (AMI) is a major cause of mortality and health system burden in Australia and other developed countries. The major source of data on the incidence and mortality of AMI is provided by routinely collected data in hospital separations or admissions, using the World Health Organizations *International Classification of Diseases* codes such as the ICD-10 (Innes et al., 1997; World Health Organization, 1992). The accuracy of such administrative codes is critical since research-based reporting, health policy decisions, and ultimately government and hospital budgets rely heavily on the sensitivity and specificity of ICD-10 codes.

However ICD-10 codes for AMI do not represent a clinical reference standard and have limited clinical validity beyond their administrative context. For instance, there are strict rules defining allocation of ICD-10 codes. Trained coders can only report diagnoses entered into the medical record by treating clinicians. These clinicians are commonly the most junior members of the clinical team, usually in training, and errors or omissions in diagnoses are rarely reviewed or corrected (Nicholls et al., 2017; Tang et al., 2017). Despite this, medical records have traditionally been the clinical reference standard against which ICD-10 codes are validated [Welk and Kwong (2017); Wiegersma et al. (2020);]. Furthermore as more than one recent review has pointed out (McCormick et al., 2014; Metcalfe et al., 2012; Rubbo et al., 2015), for practical reasons most validation studies are restricted to cases with an ICD-10 code for AMI. As a result *missed* AMI cases are not reviewed, and so the results do not/cannot provide an estimate of the omission rate or false negative rate for AMI in ICD-10 coding. Due to these limitations of the administrative record, there is growing interest in using clinical data sources to generate a comprehensive diagnostic record (Nissen et al., 2019; Rubbo et al., 2015; Spratt et al., 2017).

The primary repository of clinical data across an increasing number of healthcare services in Australia (and worldwide) is the electronic medical record (eMR). However the primary data contained in clinical records in the eMR are relatively inaccessible to clinicians, reporting agencies and researchers. For instance, persistent electrocardiographic (ECG) ST-segment elevation is a defining characteristic of STEMI-type AMI, but one recent meta-analysis of 33 validation studies found none that validated against ECG records (Rubbo et al., 2015). A second type of AMI is non-ST segment elevation myocardial infarction (NSTEMI), whose diagnosis depends upon elevation of cardiac markers documenting myocardial injury, such as troponin test results. The same meta-analysis found very few validation studies have used this information to validate NSTEMI ICD-10 codes. Thus an outstanding question is whether the eMR contains sufficient clinical detail in an accessible form to diagnose STEMI and NSTEMI.

We have recently developed and reported a new data extraction platform that interrogates a wide range of clinical sources within the eMR called SPEED-EXTRACT (Tam et al., 2020). As such, it identifies and provides bulk access to episodes-of-care that may potentially represent an AMI diagnosis. The specific sources of clinical data were: the presence of AMI-related symptoms as free-text in 1) the “Reason-for-visit” field, or 2) the presenting information from emergency department (ED) triage; 3) an order or result recorded in the eMR for a high-sensitivity cardiac troponin test (hs-troponin), 12-lead ECG, coronary angiogram, or other tests relevant to AMI; 4) placement of the patient on a cardiac pathway care plan; 5) a scanned image from a 12-lead ECG; 6) or an ICD-10 code recorded starting with I21-I25 (for a complete description of the clinical criteria see Tam et al., 2020). Consequently, SPEED-EXTRACT allows us to determine whether the eMR contains sufficient clinical detail to diagnose STEMI and NSTEMI.

The aims of this study were to 1) determine whether the eMR provided sufficient data to reliably diagnose STEMI and NSTEMI and 2) use these eMR derived diagnosis to validate ICD-10 codes for STEMI and NSTEMI. If the eMR contains insufficient clinical detail then expert chart review will not provide a primary diagnosis for each presentation. Conversely if the eMR is sufficient then we expect to see a high rate of primary diagnosis for each presentation, and importantly, a high level of inter-rater agreement for each diagnosis. If these criteria are met, then these extracted data can serve to validate ICD-10 codes for STEMI and NSTEMI with *precise* estimates of sensitivity and specificity (i.e., within ±5 percent). To our knowledge, this is one of the first studies to validate AMI against clinical data extracted directly from the eMR.

## Method

### Ethics statement

Approval for SPEED-EXTRACT and this project was obtained from Northern Sydney Local Health District (NSLHD) Human Research Ethics Committee (HREC), reference: HREC/17/HAWKE/192

### Study Population

The cohort selection process is shown in Figure 1.

**Figure 1.**
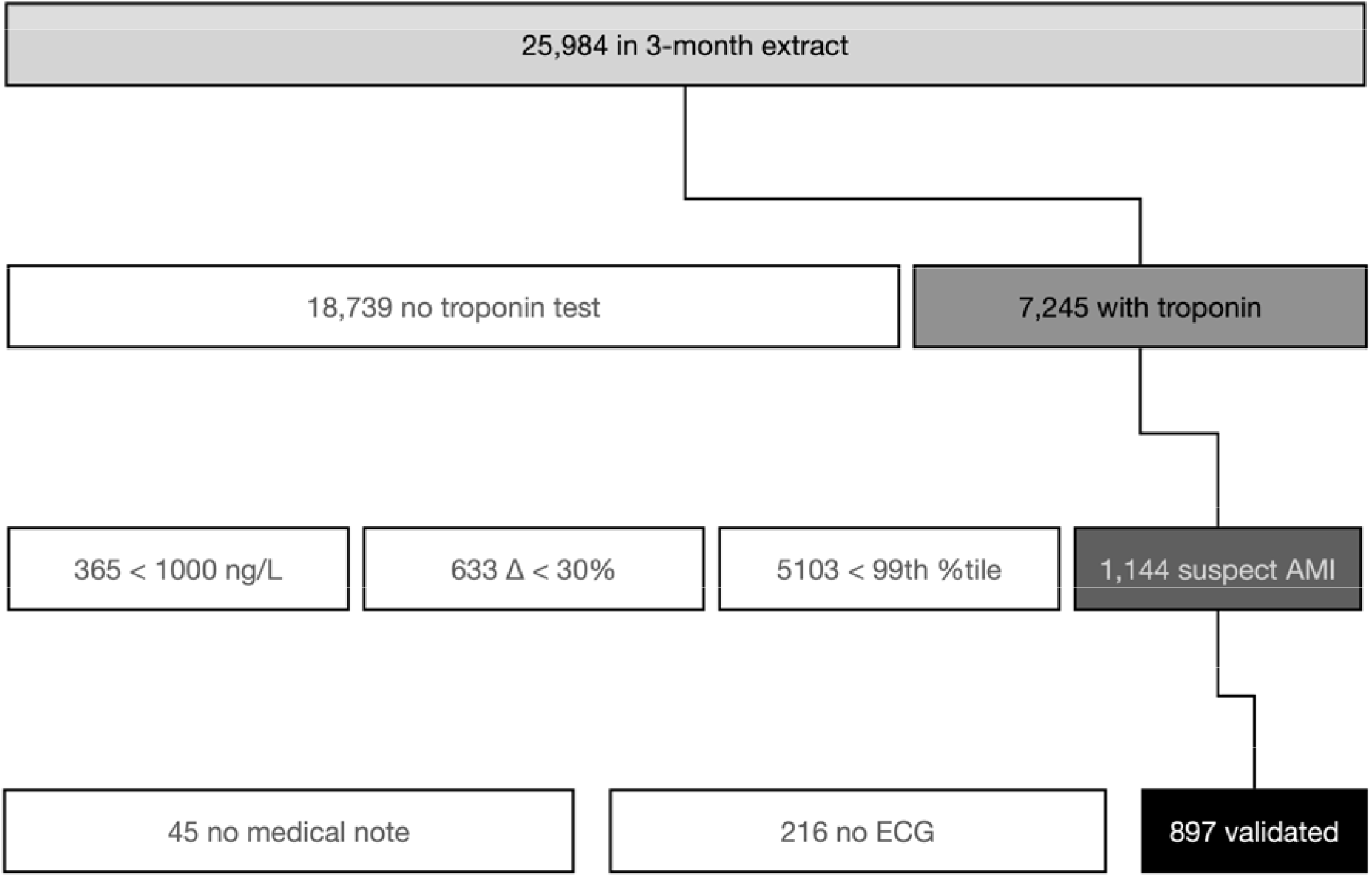
Selection of the episodes of care from the three-month eMR data extract. Note: not to scale

The SPEED-EXTRACT project represents an extract of 25,984 episodes-of-care from the eMR of Northern Sydney Local Health District and Central Coast Local Health District between April 1st to June 30th 2017. Geographically this represents two tertiary hospitals with 24-hour percutaneous coronary intervention (PCI)-capability and six referral hospitals in New South Wales, Australia. The study population for this report were adult patients within SPEED-EXTRACT who had a hs-troponin test result indicating a suspected AMI on the basis of the Universal Definition of Myocardial Infarction (UDMI) (Thygesen et al., 2019, 2012). As per the UDMI, for inclusion in the suspected AMI cohort the troponin levels of each episode had to meet the following criteria: A test result above the 99th percentile of the normal reference group according to the UDMI, and a change between two consecutive hs-troponin measurements of at least 30 percent. To capture AMI in situations in where only one troponin level was measured (a common situation encountered in clinical practice), we implemented an additional condition: A single test result greater than 1000ng/L. This resulted in *N* = 1,144 episodes-of-care in the study population with suspected AMI. Note that our study population was selected independently from the primary ICD-10 code to be validated, and so it potentially captured AMI cases which were otherwise missed by the ICD-10 code. It allowed us to provide an estimate of sensitivity, and specificity, which are both critical for valid inference when reporting AMI incidence (or prevalence).

### Study Design and Sampling

In addition to at least one hs-troponin test result, validation required at least one ECG recorded during the episode and a medical note (e.g., either the first medical note or a discharge letter, or both). Thus a further subset of episodes from the suspected AMI cohort, which met the troponin criteria (above) as well as our stringent clinical record requirements, were taken for validation; resulting in *n* = 897 episodes-of-care in the final validation sample.

As shown in Figure 1, the validated sample represents a large fraction of the *suspected AMI* cohort (78 percent, 897 / 1,144). Power analyses (see Supplementary material) indicated this strategy would produce precise estimates of the diagnostic performance of ICD-10 codes in the *suspected AMI* cohort, within ±5 percent.

### Chart review, adjudication, and validation

The *n* = 897 cases selected for validation were divided evenly between four cardiologists (SR, RH, CY, DY), such that 40 cases with similar class composition of the complete validation dataset were commonly shared among all raters to determine inter-rater reliability. This resulted in each rater reviewing approximately 233 cases. For each case, the reviewer was provided with up to the first four ECGs for the episode-of-care, the first five medical progress notes, pathology results and the angiogram report and discharge summary if available. Patient name, medical record number, and any administrative ICD-10 codes were redacted, in order to ensure blind review (and privacy). On the basis of each review, the cardiologist provided a diagnostic class from four options: STEMI, NSTEMI, unstable angina, or “other”. Importantly for our purposes, they were also required to indicate if a diagnosis could not be made if “other” was selected. Due to the low numbers of unstable angina (*n* = 10, or 1 percent of the sample), the unstable angina group was collapsed into “other” for the analysis (the slightly larger group will be referred to as “Other”) resulting in three ultimate diagnostic classes: STEMI, NSTEMI and Other.

Inter-rater agreement among the four cardiologists was determined by the percentage of cases (*n* = 40) for which there was agreement among the three diagnostic classes (STEMI, NSTEMI and Other). To adjust for the chance level of agreement among our four cardiologists, the Fleiss’ kappa was also calculated — this can be interpreted as equivalent to Cohen’s kappa for more than two raters (where agreement due to chance □= 0 and perfect agreement □= 1).

Disagreements were adjudicated by a fifth senior cardiologist (DB) and the final diagnosis was compared to the Australian Modification of ICD-10, which provides distinct codes for STEMI (I21.0, I21.1, I21.2, I21.3) and NSTEMI (I21.4) (National Centre for Classification in Health (Australia), 2004).

To assess diagnostic performance, we determined sensitivity, specificity and positive predictive values for each of the diagnostic classes. In-sample results were calculated for the validation sample (*n* = 897) using an objective Bayesian procedure, drawing k = 10,000 samples from a beta distribution for each parameter with uniform priors (Mossman and Berger, 2001). The mean and 95% credible interval of the posterior distribution is presented for each parameter. To extrapolate our results to the larger *suspected AMI* cohort (*N* = 1,144), we also simulated the experiment design with fixed test characteristics (e.g., disease prevalence in the larger sample), and the disease process was a random binomial draw accordingly. In this manner, diagnostic performance was evaluated over the wider out-of-sample distribution, from which we calculated mean and 95% CI. Using this method, the half-width of the credible interval represents the obtained uncertainty in our estimates (i.e., precision), rather than the expected uncertainty at a theoretical limit (as provided by frequentist estimates). Thus we can appropriately summarize the diagnostic performance (and its uncertainty/precision) in the validation cohort as well as the wider *suspected AMI* cohort.

## Results

### Demographics and incidence

The demographic characteristics of the validation sample are shown in Table 1 below.

**Table 1.**
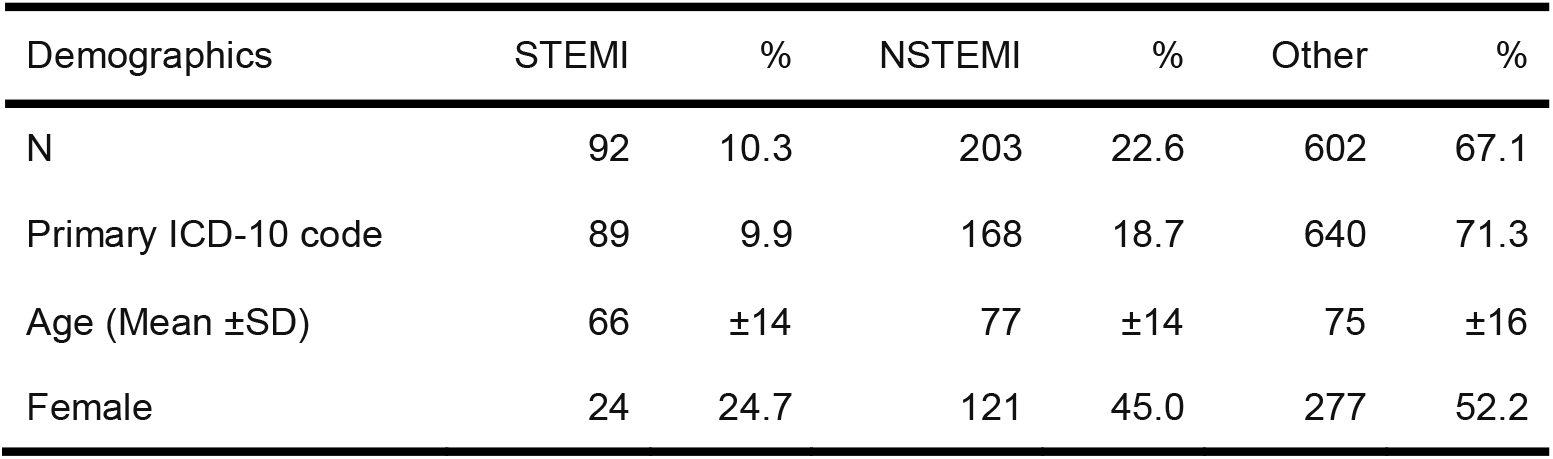
Patient characteristics in the validation cohort

The overall proportion of AMI in the validation sample was 32.9 percent (i.e., STEMI + NSTEMI). STEMI was 10.3 percent of the validation sample, while the ICD-10 coded STEMI was 9.9 percent. The ratio of NSTEMI to STEMI was approximately 2:1, consistent with previous cohorts (Chew et al., 2016; Nedkoff et al., 2017).

The remaining 247 episodes (1,144 - 897, i.e., 21.9%) from the *suspected AMI* cohort could not be validated due to missing ECG records or medical notes in the eMR. Of these missing cases, the distribution of missing cases was very similar across diagnostic classes: STEMI = 24/121 (19.8 percent); NSTEMI = 75/344 (21.8 percent); and Other = 148/679 (21.8 percent). The evenly distributed missingness across diagnostic classes indicated no diagnosis was more likely to be missing records than the other diagnoses, consistent with the MAR (missing-at-random) assumption - an important prerequisite for valid inference in the out-of-sample estimates.

### Inter-Rater Agreement

Among the three diagnostic classes: STEMI, NSTEMI, and Other (where most diagnoses are “Other”), the overall inter-rater agreement was 67.5 percent, which represents the percentage of cases from *N* = 40 for which there is agreement among the four clinician reviewers. The Fleiss’ □= 0.66 (*p* < .001), which is a moderate level of agreement among all three diagnostic classes after adjusting for chance (i.e., chance = 0). The □within each STEMI and NSTEMI subtype indicated that among the four cardiologists there were high levels agreement for the STEMI diagnosis (□= 0.79, *p* < .001), and moderate levels of agreement for the NSTEMI diagnosis (□= 0.55, *p* < .001).

### Overall Classification Results

Once the disagreements (n = 9) were adjudicated by our fifth expert (DB), the classification results for all cases in the validation sample were calculated and are shown in Table 2 below:

**Table 2.** ICD-10 (row) by clinical diagnosis (column)

|  |  | STEMI | NSTEMI | Other |
| --- | --- | --- | --- | --- |
| ICD-10 | STEMI | 81 | 4 | 4 |
|  | NSTEMI | 0 | 140 | 28 |
|  | Other | 11 | 59 | 570 |

Table 2 is a confusion matrix which summarizes the types of errors by ICD-10 that occurred in each diagnostic class. The majority of ICD-10 STEMIs were confirmed by clinical review (n = 81), although a small number of STEMI patients were miscoded as “Other” (n = 11); and none were miscoded as NSTEMI. Likewise, the majority of ICD-10 NSTEMIs were confirmed by clinical review (n = 140), and among the errors most were miscoded as “Other” (n = 59) and very few miscoded as STEMI (n = 4). Among the “Other” ICD-10 codes, most of the errors were more likely to be miscoded as NSTEMI (n = 28) than STEMI (n = 4). Thus, the pattern of errors shown in Table 2 indicate NSTEMI and “Other” were more difficult to distinguish from each other than STEMI.

There were no cases indicated as unable to diagnose by our clinical experts, confirming that within our selected validation sample there was sufficient clinical detail to provide an AMI diagnosis.

### Diagnostic performance of ICD-10 coding

The detailed *in-sample* diagnostic performance of the ICD-10 STEMI and NSTEMI coding among our *n* = 897 cases in the validation sample is shown in Table 3 below. For comparison with previous literature, we also report the result of collapsing both STEMI and NSTEMI into a single AMI class.

**Table 3.**
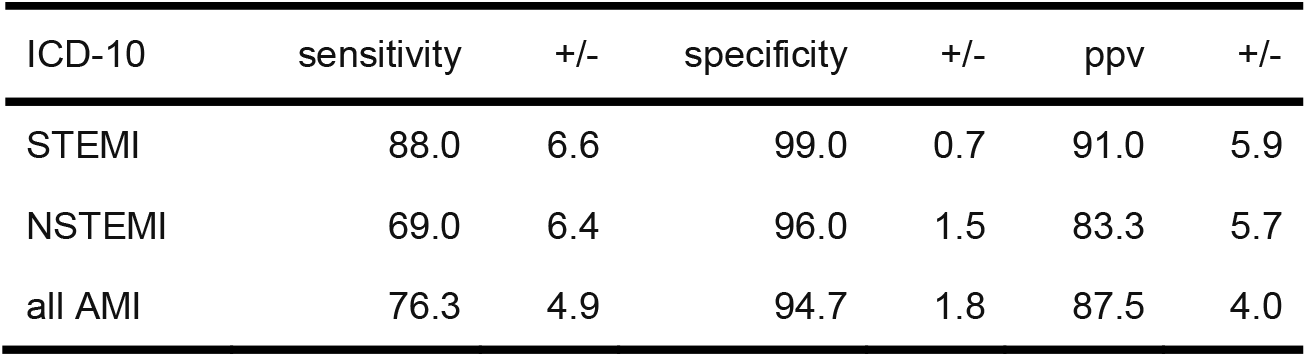
In-sample estimates ±95% CI half-widths

Table 3 shows the in-sample estimates of ICD-10 sensitivity, specificity and PPV. Generally, the specificity estimates were high (greater than 95 percent) with STEMI the highest, and with low uncertainty in each case (less than two percent in the +/- column) indicating a high level of precision. By comparison, the in-sample sensitivity estimates were lower (STEMI was highest at 88 percent) with more uncertainty (greater than 5 percent). The lowest estimate in Table 3 was NSTEMI sensitivity at 69 percent.

Only after collapsing the diagnostic groups into a single class (all AMI) did we obtain the expected level of precision (5 percent or less uncertainty) across all metrics (sensitivity, specificity, and PPV).

Note also that the low levels of ICD-10 sensitivity correspond to complementary high false negative rates (e.g., FNR_STEMI_ = 12 percent), which represent the proportion of cases missed by ICD-10 coding.

*Out-of-sample* diagnostic performance among the *suspected AMI* cohort (*N* = 1,144) (of which our validation sample represents 78 percent) is shown in Table 4 below.

**Table 4.** Out-of-sample estimates (±95% CI half-widths)

| ICD-10 | sensitivity | +/- | specificity | +/- | ppv | +/- |
| --- | --- | --- | --- | --- | --- | --- |
| STEMI | 88.1 | 6.4 | 99.0 | 0.7 | 91.0 | 6.0 |
| NSTEMI | 68.9 | 6.5 | 96.0 | 1.4 | 83.3 | 5.9 |
| all AMI | 76.3 | 4.8 | 94.7 | 1.8 | 87.6 | 4.5 |

The out-of-sample estimates for the *target_suspected AMI* cohort were very similar to the in-sample estimates from the validation cohort, with slight variations in precision of the estimates. Some variation (i.e., increased uncertainty) is expected since we are extrapolating from our validation sample, which represents 78 percent (897/114) of the *suspected AMI* cohort.

The complete posterior distribution of each parameter is shown in Figure 2.

**Figure 2.**
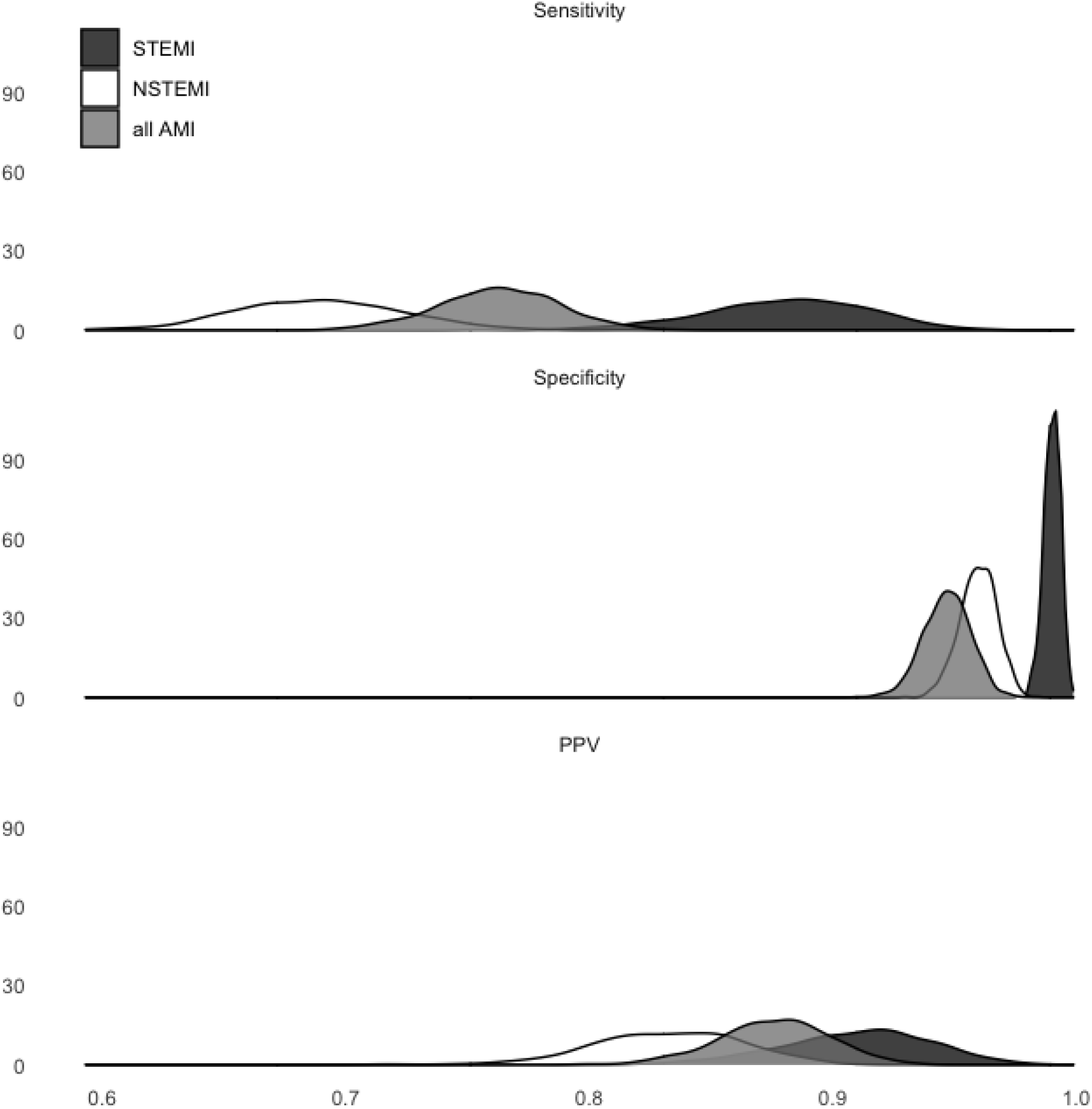
Out-of-sample distributions (density)

For each diagnostic performance metric, the mass of the STEMI distribution (dark grey) almost never overlaps the mass of the NSTEMI distribution (white). Thus, Figure 2 makes clear that the diagnostic performance of ICD-10 STEMI is almost certainly greater than ICD-10 NSTEMI for specificity, sensitivity and PPV.

## Discussion

### Principal findings

The incidence and outcomes following AMI are reportable metrics for governments and health services throughout the developed world, where they are used to evaluate performance of a healthcare system. However administrative codes for AMI (e.g., ICD-10) are an imperfect representation of the true diagnoses, and better methods are required. We used the data sources in SPEED-EXTRACT to determine whether the eMR contains sufficient clinical detail to diagnose the subcategories of AMI. Expert chart review using these data sources was able to reliably diagnose STEMI, indicated by high inter-rater agreement among our clinical experts, however the clincal detail held in the eMR was less sufficient to reliably diagnose NSTEMI, which had lower inter-rater agreement. Furthermore, using these eMR determined diagnoses as gold standards, we found ICD-10 codes identified STEMI with acceptable sensitivity and excellent specificity, but were less sensitive and marginally less specific for the diagnosis of NSTEMI.

Among the *N* = 1,144 suspected AMI cases, 78 percent (*n* = 897) had the clinical records in the eMR required for diagnosis. Validation demonstrated that among the 78 percent of cases with available records, the precision of our *in-sample* estimates of specificity and PPV were within our acceptable limit (±5%), however the precision of our sensitivity estimates were either on the boundary or exceeding the ±5% limit. Furthermore, the sensitivity, specificity, and PPV of the out-of-sample estimates of ICD-10, and their precision, did not greatly change when extrapolating to the entire cohort of suspected AMI cases (*N* = 1,144). This represents some evidence that clinical data sources within the eMR are sufficient to monitor and track AMI in a large proportion of the target population, and inferences can be made to the wider population they represent without loss of precision. Finally, the validation of ICD-10 codes demonstrated that up to a quarter of cases may be missed in administrative records. In particular, we found a 12 percent false negative rate for STEMI and a 31 percent false negative rate for NSTEMI. The high proportion of missed cases among ICD-10 codes adds weight to calls from other researchers to use routinely collected health care data for research and reporting, rather than rely on administrative data alone (Nissen et al., 2019; Welk and Kwong, 2017; Xu et al., 2020).

### Strengths and weaknesses

Previous Australian validation studies of administrative AMI codes (ICD-9) have reported sensitivity estimates in the range of 79-86 percent (Boyle and Dobson, 1995; Dobson et al., 1988), which is higher than the range we observed here (All AMI Se: 76.3±4.9 percent, see Table 3 & 4). However a relative strength of our study was validation by expert chart review of ECG records and pathology test results (e.g., hs-troponin) and medical notes, rather than the simple confirmation of a clinical diagnosis recorded in medical notes. Our independent validation against the original clinical sources is closer to the accepted gold standard, however it is rarely performed due to the historical inaccessibility of the complete medical record in the eMR (Rubbo et al., 2015). SPEED-EXTRACT shows the feasibility of using clinical sources within the eMR and these standards could be utilized in an automated pipeline to improve accessibility and accuracy of reportable outcomes such as AMI.

Our results were necessarily based on the subset of patients in our suspected AMI cohort with sufficient clinical data to make a diagnosis (897 / 1,144). Inferences from this subset to the wider suspected AMI cohort will be biased when the availability of records varies with diagnosis. For example, STEMI patients may be more likely than NSTEMI patients to have an ECG record in wider cohorts of suspected AMI patients because they were selected for ECG to obtain a diagnosis. This would result in overestimating the incidence of STEMI among the wider target cohort. Note the proportion of missing records among the AMI sub-types in our wider target cohort was approximately equivalent, and did not substantially vary with diagnosis, so we don’t think this was a factor in the present study.

### Implications

The 4th UDMI (Thygesen et al., 2019, 2012) refers to a change in cardiac troponin levels with at least one measurement in the 99th percentile to diagnose AMI. However the diagnostic agreement among our clinical experts was high for STEMI (κ = 0.786) but lower for NSTEMI (κ = 0.548). While both these agreement estimates were greater than chance (where κ = 0), the difference suggests there is either insufficient clinical detail in the eMR, or in the operational definition of the UDMI [French and White (2004); chapman2020], to reliably diagnose NSTEMI with the same level of confidence as STEMI. The current results do not let us distinguish between these two sources of uncertainty. NSTEMI, unlike STEMI, is diagnosed primarily on hs-troponin results. The 4th UDMI refers to a change in cardiac troponin levels with at least one measurement in the 99th percentile. However elevated troponin occurs in many conditions other than myocardial infarction, including in critical illnesses such as sepsis, or after chemotherapy or some renal conditions. It is for these reasons the UDMI proposes additional criteria for the classification of AMI based on pathogenesis (Alpert and Thygesen, 2016; Thygesen et al., 2019). Our clinical reference group also developed additional critiera to define our suspected AMI cohort, including a change in troponin greater than 30 percent or a single troponin measurement greater than 1000ng/L. However these criteria were still insufficient to provide clarity in the diagnosis of NSTEMI and unambiguously distinguish it from other forms of myocardial injury or “troponinemia”, as evidenced by the high proportion of cases without myocardial infarct captured in our suspected AMI sample (i.e., up to 70 percent, see also DeFilippis et al. (2019); Gronski et al. (2012)). One implication we take from this is that the operational definition of NSTEMI needs further development to provide similar levels of diagnostic confidence as STEMI. The second implication is that clinical record-keeping in the eMR must be improved to facilite accurate diagnoses for some conditions such as NSTEMI.

### Future research

The eMR is a viable source of clinical data for diagnosis of well-defined clinical conditions, such as STEMI. Clinical test results, procedures and orders stored in the eMR can be used to define and select a clinical cohort at scale, and thus reduce the reliance on administrative codes and minimize the chance of inadvertently excluding missed cases. However the application to clinical conditions which depend heavily on the clinical context in which they occur (e.g., NSTEMI) is less clear. Success in such cases will depend upon how well the clinical context can be ascertained using the information already in the eMR.

## Data Availability

Data is available from https://github.com/datarichard/SPEED-EXTRACT-validation

## Author Contributions

AS, CST, MK, MC, JG, STV, GAF, JM, & DB designed the study.

AS, CST, MK, SR, RH, CY, DZY, & DB helped carry out the study.

AS, RWM, CST, MC, MK, JG, STV, JM, & DB interpreted the results.

AS, RWM, CST, JG, STV, GAF, & DB wrote the paper.

AS, CST, MC, JG & JM secured funding for the project.

## Data sharing Statement

All deidentified data used in this study is available from `insert URL here.`

