## Supplementary material for "Validation of acute myocardial infarction (AMI) in electronic medical records: the SPEED-EXTRACT Study": validation-supplement.html

Date rendered: 08 December, 2020

---

#### Supplementary Materials

###### Clinical data availability

###### Table S1. Availability of clinical records in the validation sample

| Records | N = 8971 |
| --- | --- |
| No. ECGs | 6 (4, 9) |
| Angiogram present | 181 (20%) |
| Max Troponin value (ng/L) | 309 (68, 2,724) |
| No. medical notes |  |
| 1 | 9 (1.0%) |
| 2 | 38 (4.2%) |
| 3 | 56 (6.2%) |
| 4 | 80 (8.9%) |
| 5 | 573 (64%) |
| 6+ | 141 (16%) |
|  |  |
| --- | --- |
| 1Statistics presented: Median (IQR); n (%) | |

The remaining 247 episodes (1,144 - 897) from the *suspected AMI* cohort could not be validated due to missing ECG records or medical notes in the eMR. Of these missing cases, the distribution of ICD-10 classes were very similar: 24/121 STEMI (19.8%), 75/344 NSTEMI (21.8%), and 148/679 “Other” (21.8%), consistent with the missing cases missing-at-random (MAR).

###### Power analysis by simulation

For simulation we considered the test characteristics as fixed or given, and then simulated the disease status and test performance as a random process according to those parameters. By doing this we can appropriately summarise the test performance (and its uncertainty or precision) in a variety of scenarios, such as over a range of values for disease prevalence or in larger samples.

The basic parameters are given as follows:

\[
\begin{array}{c|ccc}
& D & \bar{D} & \\ \hline
T & TP & FP& n\_t\\
\bar{T} & FN & TN & n-n\_t\\
& n\_d & n-n\_d
\end{array}
\]

Where \(n\_t\) is the number of cases who tested positive, and \(n\_d\) is the true number of cases with the disease.

Then \(TN\) and the \(FN\) are binomial probability models where:

\[
n\_d \sim Binomial(\pi, n)
\] \[
TP \sim Binomial(\theta\_{Se}, n\_d)
\] \[
FN \sim Binomial(\theta\_{Sp}, n - n\_d)
\]

Where \(\pi\) is the prevalence of the disease, and \(\theta\) is the sensitivity and specificity, respectively.

#### Supplementary Results

###### Power analysis

The effect of increasing the sample size and disease prevalence on the precision of our estimates is shown below. Prevalence (\(\pi\) = 0.1) and sample size (\(\n\) = 800) were held constant otherwise.

Increasing the sample size from *n* = 400 to *n* = 1000 improved the precision of our estimates for sensitivity (Se) and specificity (Sp) (i.e., reduced uncertainty). The precision of specificity was well-below our minimum threshold (5%) for each level of Sp tested. However the precision of sensitivity only reached the 5 percent threshold for high values of Se and when the sample size was *n* > 800.

At a fixed sample size (*n* = 800), increasing the disease prevalence from \(\pi\) = 0.05 to 0.2 improved the precision of our estimates for senstivity and specificity (i.e., reduced uncertainty). The precision of specificity was well-below our minimum threshold (5%) for each level of Sp tested. However the precision of sensitivity only reached the 5 percent threshold for high values of Se and \(\pi\) > 0.1.

### Do not include

###### Sensitivity analysis

Our *suspected AMI* cohort was mostly comprised of cases that were neither STEMI nor NSTEMI (67.1 percent “Other”, see Table 1). While this is in-line with previous reports indicating myocardial injury may comprise over 70 percent of a cohort meeting UDMI criteria [@defilippis2019; @gronski2012], we were concerned that our results may be unduly influenced by our reliance on troponin test results.

Due to such concerns, we conducted a sensitivity analysis of the suspected AMI cohort which was further restricted to cases with key words such as “chest pain” in the Reason for visit field or Presenting problem field of SPEED-EXTRACT. The complete list of keywords is provided in Supplementary materials.

To define a stricter clinical cohort, the following keywords were searched in the Reason for visit field or Presenting problem field of SPEED-EXTRACT.

###### Keywords

*‘chest pain’, ‘pain chest’,‘pain, chest’,‘pain-chest’,‘shortness of breath’, ‘sob’, ‘dizziness’, ‘vomiting’, ‘syncope’, ‘syncopal’, ‘presyncope’, ‘weakness’, ‘nausea’, ‘unwell’, ‘loc’,‘cardiac arrest’, ‘nstemi’, ‘stemi’, ‘angio’, ‘angiogram’, ‘cor/angio’, ‘palpitations’, ‘salami’, ‘cabg’, ‘ami’, ‘coronary artery bypass graft’, ‘etami’, ‘heart attack’, ‘cath lab’, ‘cath’, ‘ohca’, ‘out of hospital arrest’, ‘out of hospital cardiac arrest’, ‘stent’, ‘fatigue’, ‘weakness’, ‘ventricular tachycardia’, ‘ventricular fibrillation’, ‘vt’, ‘vf’, ‘dyspnoea’, ‘chest tightness’*.

###### Results for sensitivity analysis

After including the Reason for visit information along with troponin result in a stricter cohort definition, the number of cases in the validation sample decreased from *n* = 897 to *n* = 629. The resulting numbers and incidence of AMI in the stricter clinical cohort are shown below in Table S2.

###### Table S2. Frequency and incidence of AMI in clinical cohort

|  | STEMI | % | NSTEMI | % | Other | % |
| --- | --- | --- | --- | --- | --- | --- |
| Diagnosis | 84 | 13.4 | 167 | 26.6 | 378 | 60.1 |
| ICD10 | 85 | 13.5 | 148 | 23.5 | 396 | 63.0 |

Table S2 shows the proportion of the sample comprised of AMI increased slightly to 40 percent, representing approximately a ten point increase and indicating our stricter cohort definition was partially successful in excluding non-AMI cases from the validation sample.

The classification results in the stricter clinical cohort, along with diagnostic performance of sensitivity, specificity and PPV are shown in Tables S3 and S4.

###### Table S3. ICD-10 (row) by diagnosis (column)

|  |  | STEMI | NSTEMI | Other |
| --- | --- | --- | --- | --- |
| ICD-10 | STEMI | 78 | 3 | 4 |
|  | NSTEMI | 0 | 127 | 21 |
|  | Other | 6 | 37 | 353 |

###### Table S4. Clinical in-sample estimates (±95% CI half-widths)

| ICD-10 | sensitivity | +/- | specificity | +/- | ppv | +/- |
| --- | --- | --- | --- | --- | --- | --- |
| STEMI | 0.93 | 0.05 | 0.99 | 0.01 | 0.92 | 0.06 |
| NSTEMI | 0.76 | 0.06 | 0.95 | 0.02 | 0.86 | 0.06 |
| all AMI | 0.83 | 0.05 | 0.93 | 0.03 | 0.89 | 0.04 |

Generally, a stricter clinical cohort definition slightly improved the classification results relative to our original cohort. By including the presenting problem in our cohort definition, we reduced the incidence of non-AMI cases in the validation sample by around 10 percent. We also observed small increases in the sensitivity and positive predictive value of AMI and each sub-type, attributable to reductions in the number of false positive cases (false negative cases were also reduced slightly). For instance, false positive STEMI cases were almost halved (from *n* = 11 to 6), which increased the sensitivity of ICD-10 STEMI to greater than 90 percent. However the diagnostic sensitivity for NSTEMI was approximately 17 points below STEMI, still suggesting NSTEMI was more difficult to distinguish than STEMI.

###### Error analysis of false negative NSTEMI

The 10 percent lower sensitivity scores for NSTEMI were due to the high rate of missed NSTEMI. What ICD-10 diagnoses occur in these missed NSTEMI?

Top 10 ICD-10 diagnoses in missed NSTEMI

| SOURCE\_STRING | n | % |
| --- | --- | --- |
| Acute subendocardial myocardial infarction | 32 | 103 |
| Hypertension | 25 | 81 |
| Personal history of tobacco use disorder | 20 | 65 |
| Essential (primary) hypertension | 18 | 58 |
| Place of occurrence, health service area | 15 | 48 |
| Atherosclerotic heart disease, of native coronary artery | 12 | 39 |
| Acute kidney failure, unspecified | 11 | 35 |
| Atrial fibrillation and atrial flutter, unspecified | 11 | 35 |
| Congestive heart failure | 11 | 35 |
| Hypotension, unspecified | 11 | 35 |

Top 10 ICD-10 diagnoses in true NSTEMI

| SOURCE\_STRING | n | % |
| --- | --- | --- |
| Acute subendocardial myocardial infarction | 140 | 100 |
| Hypertension | 69 | 49 |
| Atherosclerotic heart disease, of native coronary artery | 65 | 46 |
| Personal history of tobacco use disorder | 48 | 34 |
| Essential (primary) hypertension | 28 | 20 |
| Type 2 diabetes mellitus without complication | 22 | 16 |
| Presence of coronary angioplasty implant and graft | 19 | 14 |
| Tobacco use, current | 18 | 13 |
| Hypotension, unspecified | 17 | 12 |
| Place of occurrence, health service area | 17 | 12 |

Top 10 differences

| ICD10 diagnosis | p\_true | p\_missed | delta |
| --- | --- | --- | --- |
| Essential (primary) hypertension | 20 | 58 | 38 |
| Hypertension | 49 | 81 | 32 |
| Personal history of tobacco use disorder | 34 | 65 | 31 |
| Pneumonia, unspecified | 4 | 32 | 28 |
| Unstable angina | 8 | 35 | 27 |
| Atrial fibrillation and atrial flutter, unspecified | 9 | 35 | 26 |
| Congestive heart failure | 9 | 35 | 26 |
| Acute kidney failure, unspecified | 10 | 35 | 25 |
| Hypotension, unspecified | 12 | 35 | 23 |
| Type 2 diabetes mellitus with established diabetic nephropathy | 6 | 29 | 23 |
| Arthritis and osteoarthritis | 10 | 32 | 22 |
| Type 2 diabetes mellitus with other specified kidney complication | 1 | 23 | 22 |
| Ischaemic heart disease | 10 | 29 | 19 |
| Respiratory failure unspecified, type II | 1 | 19 | 18 |
| Disorders of magnesium metabolism | 6 | 23 | 17 |
| Urinary tract infection, site not specified | 6 | 23 | 17 |
| Type 2 diabetes mellitus without complication | 16 | 32 | 16 |
| Left ventricular failure | 7 | 23 | 16 |
| Personal history of long-term (current) use of other medicaments, insulin | 8 | 23 | 15 |
| Hypokalaemia | 4 | 19 | 15 |
